# Improving diagnosis of non-malarial fevers in Senegal: *Borrelia* and the contribution of tick-borne bacteria

**DOI:** 10.1101/2023.08.24.23294564

**Authors:** Zoë C. Levine, Aita Sene, Winnie Mkandawire, Awa B. Deme, Tolla Ndiaye, Mouhamad Sy, Amy Gaye, Younouss Diedhiou, Amadou M. Mbaye, Ibrahima Ndiaye, Jules Gomis, Médoune Ndiop, Doudou Sene, Marietou Faye Paye, Bronwyn MacInnis, Stephen F. Schaffner, Daniel J. Park, Aida S. Badiane, Andres Colubri, Mouhamadou Ndiaye, Ngayo Sy, Pardis C. Sabeti, Daouda Ndiaye, Katherine J. Siddle

## Abstract

The worldwide decline in malaria incidence is revealing the extensive burden of non-malarial febrile illness (NMFI), which remains poorly understood and difficult to diagnose. To characterize NMFI in Senegal, we collected venous blood and clinical metadata from febrile patients and healthy controls in a low malaria burden area. Using 16S and unbiased sequencing, we detected viral, bacterial, or eukaryotic pathogens in 29% of NMFI cases. Bacteria were the most common, with relapsing fever *Borrelia* and spotted fever *Rickettsia* found in 15% and 3.7% of cases, respectively. Four viral pathogens were found in a total of 7 febrile cases (3.5%). Sequencing also detected undiagnosed *Plasmodium*, including one putative *P. ovale* infection. We developed a logistic regression model to distinguish *Borrelia* from NMFIs with similar presentation based on symptoms and vital signs. These results highlight the challenge and importance of improved diagnostics, especially for *Borrelia*, to support diagnosis and surveillance.

## Introduction

Febrile illness is a significant cause of morbidity and mortality in West Africa. While malaria remains the most common single pathogen causing febrile illness, its incidence has decreased sharply in the last two decades; in Senegal, for example, control measures decreased malaria incidence from 122 per 1000 in 2006 to 59 per 1000 in 2021^1,2^. As the malaria burden has decreased, the importance of non-malarial febrile illness (NMFI) has become more apparent. Unlike malaria, however, many of the pathogens causing NMFI are not the target of robust surveillance programs, and rapid diagnostic tests (RDTs), the backbone of both clinical care and surveillance, are not available^3^.

A broad range of pathogens across kingdoms can cause NMFI, but surveillance studies often focus on one or a few pathogens and therefore are limited in their ability to detect all causes of disease. Causal pathogens include arboviruses, such as Dengue virus and Chikungunya virus, which cause both sporadic febrile illness and outbreaks in Senegal^4,5^. Bacterial pathogens are a common cause of severe illness requiring hospitalization, including several vaccine-preventable illnesses such as *Streptococcus pneumoniae* and *Neisseria meningitidis*^6^ and bacterial zoonoses are increasingly recognized as a common cause of ambulatory febrile illness, including tick and louse-borne relapsing fevers, spotted fever, Q fever, Leptospirosis and Brucellosis^6–8^. While fungal infections appear less common, *Candida* and *Cryptococcus* have been reported across West Africa, with cryptococcal disease being particularly common in HIV-positive patients^6^. Non-malarial parasitic diseases including trypanosomiasis and leishmaniasis are also common^6^.

In the absence of comprehensive surveillance efforts that capture multiple pathogen types, appropriate public health interventions are hindered by our limited understanding of the full landscape of common causes of NMFI in the region. Unbiased sequencing, also known as metagenomic sequencing (mNGS), is a powerful tool for detection of microbial nucleic acids in clinical samples without *a priori* knowledge of a pathogen^9^ and is increasingly used for surveillance in regions at high risk of emerging and reemerging disease^10–13^. However, the low abundance of pathogen nucleic acids relative to host and commensal microbial material requires deep sequencing and can lead to false conclusions^14,15^. Unbiased approaches are complemented by more targeted strategies, such as 16S sequencing for bacterial pathogens^16^, that specifically amplify pathogen nucleic acids reducing required sequencing depth and simplifying interpretation. This technique has been applied to detect bacterial bloodstream infections^17,18^ and tick-borne bacterial illness^19^.

This study aimed to determine and characterize the major causes of NMFI in Thiès, Senegal, a peri-urban community with overall low malaria incidence where malaria transmission dynamics have been deeply characterized^20^. We collected plasma samples from a cohort of acutely febrile patients presenting to the Service de Lutte Anti Parasitaire (SLAP) clinic and healthy controls from the surrounding neighborhoods during the dry and rainy seasons of 2018 and 2019. We aimed to provide insights on clinical presentations of NMFI that can guide providers at the point of care and genomic characterization to inform design of new detection tools for clinical diagnosis and public health surveillance.

## Results

### Broad characterization of NMFI reveals viral, bacterial, and eukaryotic pathogens

We first comprehensively characterized the pathogens in plasma samples collected from acutely febrile patients suspected of malaria and healthy controls across the dry (febrile: n = 100, healthy: n = 54) and rainy (febrile: n = 104, healthy: n = 50) seasons in 2019 (Fig. 1A, Supplementary Fig. 1B). All febrile patients received a malaria RDT; 39.4% (41/104) of patients tested positive for malaria in the wet season and no cases were detected in the dry season. Febrile cases were roughly equally split between children and adults (50.4% >= 18yrs). Age distribution was similar (p = 0.98, Mann-Whitney, two-sided) across case and control groups, but there were more male febrile cases (Male: n = 110, Female: n = 91) and more female controls (Male: n= 47, Female: n = 57, p = 0.15 Fisher exact two-sided, Supplementary Fig. 1A).

**Figure 1:**
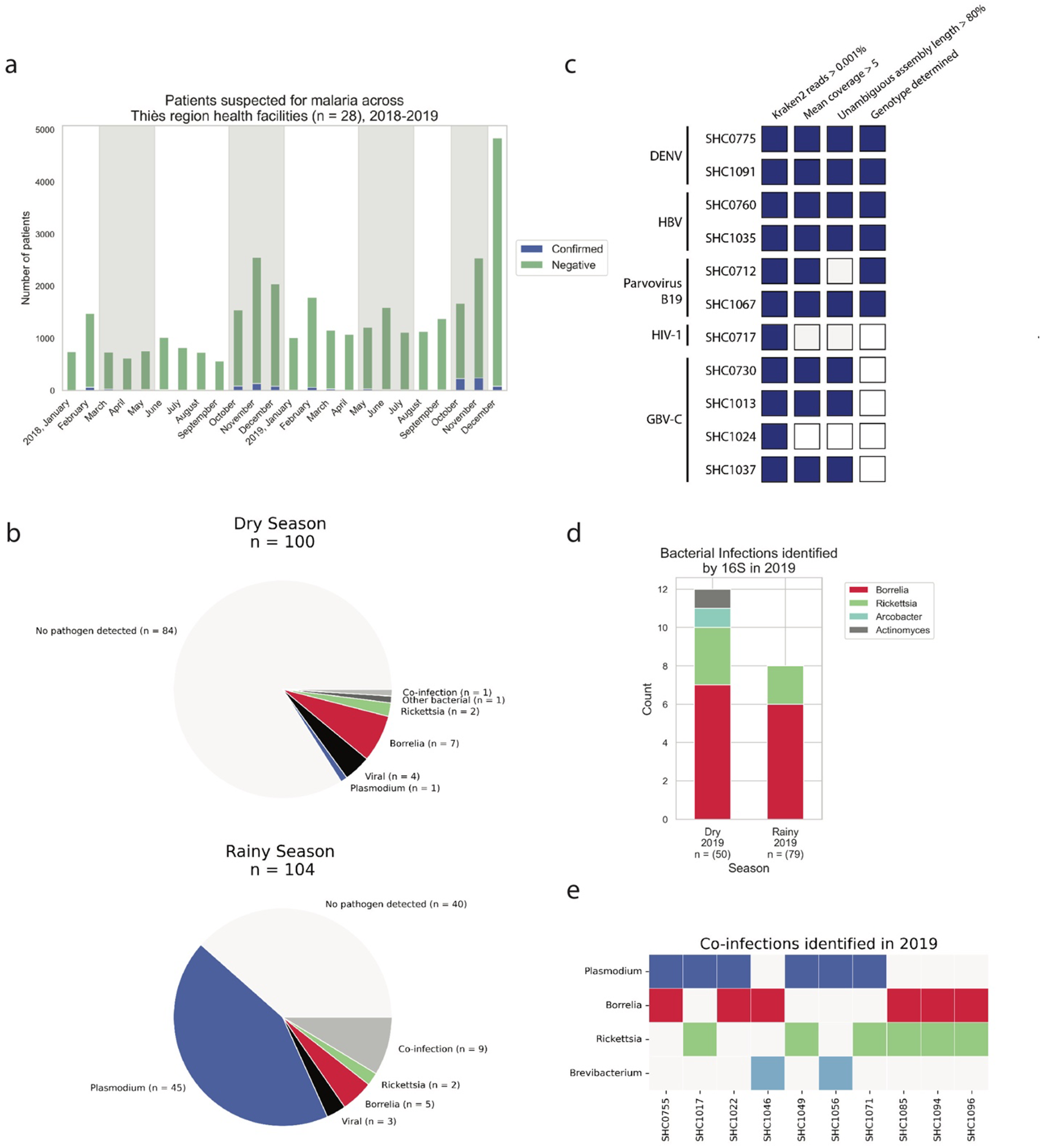
Overview of burden of malarial and NMFI in Thiès, Senegal and NMFI pathogens identified in 2019 **a.** Patients suspected of malaria across 28 health facilities in the Thiès region (data provided by Programme National de lutte contre le Paludisme); gray shading indicates periods of sample collection for this study, **b.** Types of pathogens detected in febrile patients in dry and rainy seasons in 2019. Plasmodium detected by RDT or RNA-mNGS, bacterial infections detected by 16S amplicon sequencing, and viral infections detected by RNA-mNGS. **c.** Viral infections detected by RNA-mNGS **d.** Bacterial infections detected by 16S sequencing and **e.** Co-infections detected in febrile patients from 2019.

We performed broad detection of viral, bacterial, and eukaryotic pathogens by sequencing. To detect viral pathogens, we performed RNA-mNGS. We also evaluated the ability of RNA-mNGS to detect non-viral pathogens, including malaria and fungi. To detect bacterial infections, we sequenced the v1-2 region of the 16S rRNA gene, which permitted us to classify the bacterial taxa present in samples with high bacterial load [see methods]. We detected at least one pathogen in 24% (39/163) of RDT-negative acutely febrile patients and co-infections in 7% (3/41) of RDT-positive acutely febrile patients (Fig. 1B). The most common febrile pathogen was *Borrelia*, which was found across the dry (n = 8) and rainy (n = 10) seasons and in both RDT-negative (n = 17) and RDT-positive (n = 1) patients. We also detected bacterial infections with *Rickettsia, Arcobacter, Actinomyces* and *Brevibacterium* (Fig. 1D). Bacterial pathogens found as co-infections included three *Borrelia/Rickettsia* co-infections, two *Borrelia/Plasmodium* co-infections, and three *Plasmodium/Rickettsia* co-infections (Fig. 1E).

We identified four known vertebrate viruses, Dengue virus (DENV, n = 2), Hepatitis B virus (HBV, n = 2), Parvovirus B19 (n = 2), and Human immunodeficiency virus 1 (HIV-1, n = 1) as well as a human virus not currently believed to cause disease, Pegivirus C (GBV-C, n = 4) (Fig. 1C). Four febrile patients and one healthy control exhibited a high proportion of fungal RNA reads in the plasma, as compared to healthy controls (> 99th percentile reads/million raw reads (rpm) for healthy controls), but these reads did not map to any specific fungal pathogen (Supplementary Fig. 8A). We did not detect any vertebrate viruses in healthy controls, but we did detect *Borrelia* (n = 1/35 samples sequenced) and *Rickettsia* (n = 1/35).

### RNA-mNGS identifies viral pathogens known to circulate in Senegal

We next considered the genetic diversity of detected viral pathogens to determine their relationship to other circulating strains in West Africa. Phylogenetic analysis of complete DENV, HBV and Parvovirus B-19 genomes indicated that in all three cases, the two isolates were different genotypes, and thus not closely related (Supplementary Table 1). Notably, there was a DENV outbreak across Senegal, including Thiès, in 2018. Whole genome phylogenetic analysis revealed the closest relatives to the 2019 DENV3 isolate from this study were DENV3 genomes from patients presenting to the SLAP clinic during that outbreak^4^ (Supplementary Fig. 2A). Conversely, the DENV1 genome in this study was more closely related to other isolates from West Africa than to the DENV1 isolates from the 2018 outbreak (Supplementary Fig. 2A). The HIV-1 isolate genome could not be assembled and genotyped due to low read count (mean coverage 0.2), but reads mapped across the reference genome including in the *gag, pol,* and *env* genes.

While GBV-C is known to infect asymptomatic individuals across the world, we only detected GBV-C in febrile patients. The prevalence of GBV-C has not been well studied in Senegal; we found infection in only 1.3% (4/307) of plasma samples sequenced, lower than the 4-11% reported in blood-donor surveys from Sub-Saharan Africa^21,22^. All four isolates were clustered with previously sequenced human isolates from Sierra Leone, Uganda and Cameroon and belonged to Genotype I (Supplementary Fig. 2B), the most commonly circulating GBV-C genotype in West Africa^23^.

In order to identify divergent viral species missed at the read level, we performed a translated nucleic acid search of *de novo* contigs which identified RNA-dependent RNA Polymerase (RdRp) sequences for two candidate novel viruses, one in the *Naranviridae* family and one in the *Reoviridae* family. We detected a *Reoviridae* RdRp sequence across 18 febrile patients, 12 healthy controls, and 4 non-template controls, suggesting a likely contaminant (Supplementary Fig. 3). However, given that *Reoviruses* have been isolated from ill patients, we further investigated these sequences^24^. The *Reovirus* RdRp sequences from this study clustered together, but were distant from mammalian *Orthoreoviruses*. Given the distance from mammalian *Orthoreovirus* species and presence in non-template controls, these sequences were likely from a contaminated reagent rather than true human infections. We also identified a *Narnaviridae* RdRp sequence across 5 febrile patients and 1 healthy control (Supplementary Fig. 4), none of whom had any other identifiable pathogen. Because viruses in the *Narnaviridae* family, which infect plants and fungi, have not been reported as human pathogens and we detected this novel *Narnavirus* species in both febrile cases and a healthy control, the role of this virus in disease, if any, is unclear.

### Unbiased RNA sequencing detects *Plasmodium* cases missed by RDT

If RNA-mNGS is to be employed as a tool for diagnosis of febrile illness in malaria endemic regions, we also need to understand the ability of RNA-mNGS to detect malaria as compared to clinical diagnostics. We know that RDTs have limited sensitivity, detect only certain *Plasmodium* species, and are susceptible to false negatives in genetically diverse parasites, so we evaluated our RNA-mNGS reads for evidence of *Plasmodium* infections.

RNA-mNGS detected abundant *Plasmodium* nucleic acids (>550 rpm) in 6.8% (11/160) of RDT negative acutely febrile patients, suggesting these were false negatives. Among RDT positive patients, there was a wide range in the abundance of *Plasmodium* reads (mean 1.7x10∧4 rpm, range 1.5x10∧2 – 2.6x10∧5 rpm) (Supplementary Fig. 5A). Overall, 63% of RDT positive patients (26/41) and 77% of smear positive patients (17/22) were also positive by unbiased RNA sequencing (>550 rpm) (Supplementary Fig. 5C). In patients with detectable parasitemia by blood smear examination, *Plasmodium* reads did not correlate strongly with parasite density (Pearson’s R = 0.49, Supplementary Fig. 5B).

In order to determine why 11 samples were negative by RDT but positive by RNA-mNGS, we first determined whether any samples belonged to species other than *Plasmodium falciparum,* the target species of the RDTs (Bioline Malaria Ag P.f., Abbot). As previous work has demonstrated Kraken2 is inaccurate for species-level classification of parasites^25^, we used DIAMOND to identify one sample with a contig (771 bp) that matched perfectly (100% nucleotide identity, 100% coverage) to *Plasmodium ovale* cytochrome oxidase subunit 1 (Accession: KP050416.1), suggesting a likely *P. ovale* infection. Additionally, we assessed expression levels of histidine-rich protein 2 (HRP-2), the target antigen of the RDTs used in this study. Deletions in *pfhrp2* gene have been detected in Senegal and demonstrated to cause decreased antigen expression and consequently false negative RDT results^26,27^. The number of RNA-mNGS rpm aligned to *pfhrp2* was significantly different between patients who were RDT positive and mNGS positive (RDT+/mNGS+) and patients who were RDT negative but *Plasmodium* RNA-mNGS positive (RDT-/mNGS+) (Supplementary Fig. 5D, Mann Whitney two-sided, p = 0.038). However, given the overlap in expression levels between the two groups and the low frequency of *pfhrp2* deletion in Senegal, previously estimated at 2.4%^27^, *pfhrp2* deletion likely explains only some of the false negative RDTs.

### 16S sequencing confirms a high burden of Relapsing Fever Borrelia and Spotted Fever Rickettsia

As bacteria were the most common cause of NMFI in our 2019 cohort, we extended our 16S sequencing to detect bacterial infections across the complete cohort from 2018-2019, and further investigated the species causing disease. Among all high bacterial load samples that underwent 16S sequencing [see methods], we detected *Borrelia* in 15.5% (33/213) and *Rickettsia* in 5.1% (9/213) of febrile patients. These arthropod-borne bacterial pathogens are known to circulate in Senegal and cause febrile illness (Fig. 1D, Supplementary Fig. 6C)^8^. 16S sequencing also identified other bacterial genera containing known pathogens, including *Bacteroides, Actinomyces, Brevibacterium, Arcobacter* and *Veillonella* across both years (Supplementary Fig. 6C). However, given the limited taxonomic resolution of v1-2 16S sequencing, we were unable to determine whether these sequences represented pathogenic species or harmless commensals.

Phylogenetic clustering of *Borrelia* v1-2 16S sequences showed that all *Borrelia* sequences from this study were similar to each other and fell within the Relapsing Fever (RF) group (Supplementary Fig. 7A). RF *Borrelia* circulate worldwide and are known to cause febrile illness in Senegal^28^. All the *Rickettsial* v1-2 16S rRNA gene sequences clustered within the Spotted Fever group (Supplementary Fig. 7B). Spotted fever *Rickettsia,* including *R. felis*^29^ and *R. africae*^8^ have been previously detected in febrile Senegalese patients. We did not detect any Typhus group *Rickettsia* in our cohort, though *R. typhi* has been reported in an immigrant traveling from Senegal^30^.

A subset of 6 *Borrelia* isolates were typed by amplicon sequencing of the 16S-23S intergenic spacer (IGS)^31^. All typed isolates were *Borrelia crocidurae* (Fig. 2C). Isolates from this study were more similar to previously characterized isolates from humans and *Orthnithodoros sonrai* in Southern Senegal (unpublished) and *Orthnithodoros sonrai* ticks in Mali^32^ than isolates from *Orthnithodoros erraticus* in Tunisia^33^ and Morocco^34^ (Fig. 2C).

**Figure 2:**
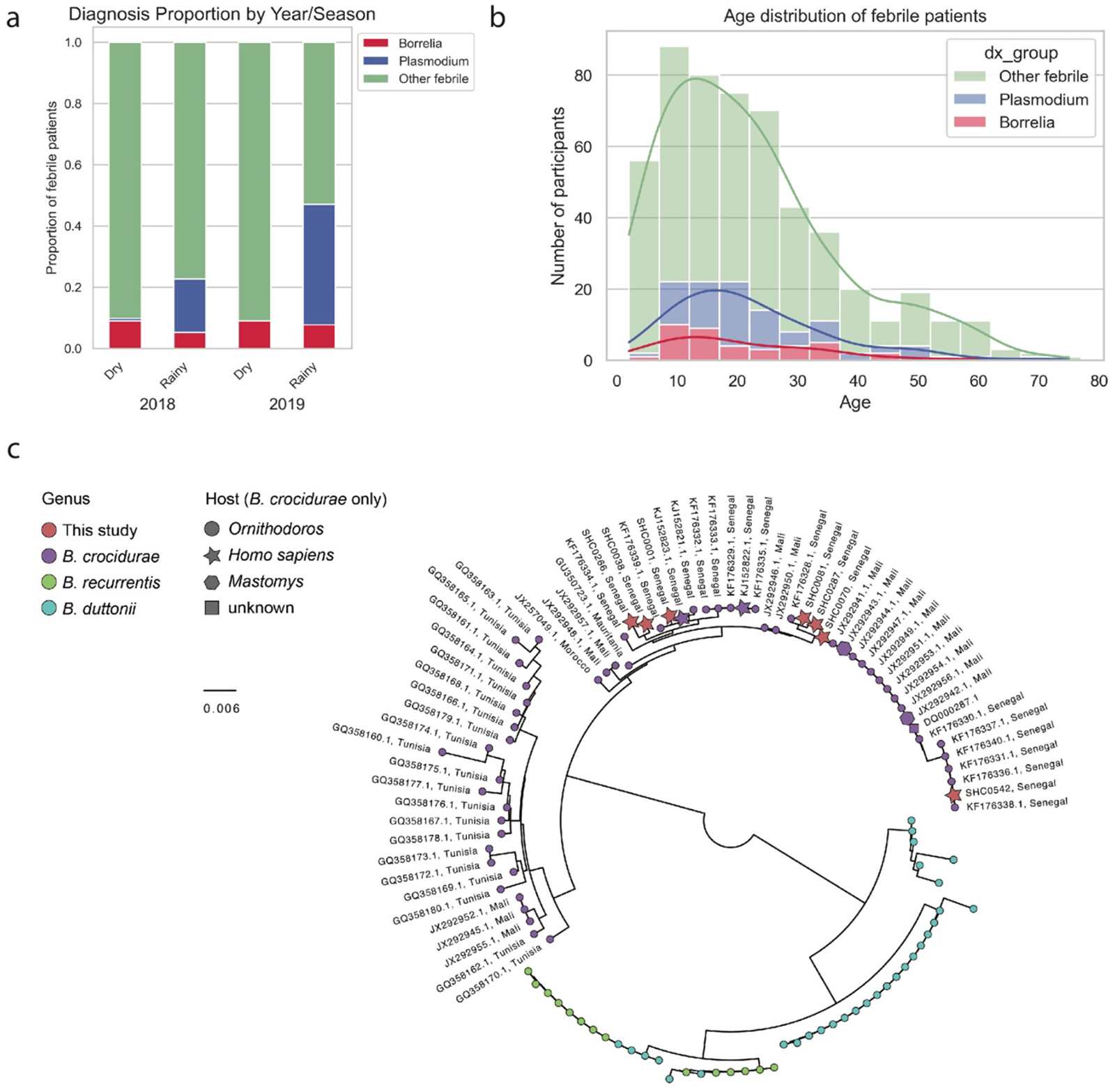
Characterization of qPCR-confirmed *8orrelia* and ROT-confirmed *P/asmodium* across both years **a.** 8orre/ia, P/asmo­ *dium,* and non-8orre/ia NMFI (Other febrile) incidence across dry and rainy seasons in 2018 and 2019. **b.** Age distribution of *8orre/ia, P/asmodium,* and other febrile patients. c. Maximum likelihood phylogenetic tree (midpoint rooted) of IGS sequences from this study (red) in the context of available reference sequences for 8. *crocidurae* (purple), 8. *duttonii* (cyan}, and 8. *recur­ rentis* (green). For 8. *crocidurae* isolates, location of collection indicated in label and host species indicated with node shape as available.

### 16S sequencing enables detection of low-titer infections

We assessed the ability of RNA-mNGS to detect bacterial infections, based on samples that underwent both sequencing methods. *Borrelia* RNA-mNGS reads were detected in 68% (13/19) of 16S positive samples. Conversely, *Rickettsia* RNA-mNGS reads were not detected in any *Rickettsia* positive samples. Acute RF *Borrelia* infection is known to cause high titers of bacteria in the blood during febrile episodes, while *Rickettsia* is an obligate intraerythrocytic pathogen and therefore has low titers in the plasma. To assess the extent to which plasma titers impacted the sensitivity of RNA-mNGS to tick-borne bacterial pathogens, we quantified bacterial abundance by the percent of total v1-2 16S sequences from a given sample classified as *Borrelia* or *Rickettsia,* respectively^19^. We observed *Borrelia* abundances ranging from 5-98%, while *Rickettsia* abundances were lower, ranging from 5% to 16%. RNA-mNGS only detected *Borrelia* in samples with abundance >20% (Supplementary Fig. 7F). Taken together, these data suggest that RNA-mNGS can detect high titer bacterial pathogens but will miss less abundant species.

‘ We compared the sensitivity of our 16S sequencing to qPCR and Giemsa stained blood smear examination, the current gold standard for clinical diagnosis of RF *Borrelia*^35^. We examined smears from a subset of RF *Borrelia* patients. 28.6% (4/14) of examined smears were positive; all 4 positive patients had a high RF *Borrelia* load (>= 90%, Mann-Whitney two-sided p = 0.002 compared to 16S positive/smear negative) (Supplementary Fig. 6B, D). Although blood smear examination is the most widely used diagnostic, qPCR diagnostics are known to have higher sensitivity^35,36^. Using a pan-*Borrelia* qPCR assay previously used for detection of *Borrelia* in patient blood samples^37^ 79% (27/34) of *Borrelia* samples were confirmed by qPCR, while 6 samples that were negative by 16S sequencing were positive by qPCR. Bacterial load measured by qPCR and 16S abundance correlated well (Pearson’s R = 0.72, Supplementary Fig. 6G). While 16S sequencing was done only on a subset of samples, pan-*Borrelia* qPCR was performed on all febrile cases and healthy controls in 2019 and all febrile cases in 2018. An additional 5 cases of *Borrelia* were identified in febrile patients by qPCR in samples that did not undergo 16S sequencing (Supplementary Fig. 7E).

### *Borrelia* infection presents similarly to other febrile illness but can be distinguished by key features

Given the limited sensitivity of available smear-based diagnostics compared to molecular methods, we sought to assess the clinical syndrome associated with qPCR-confirmed *Borrelia* infections, compared to RDT-confirmed malaria and non-*Borrelia* NMFI (“other febrile”), to guide differential diagnosis at the point of care. *Borrelia* infections occurred in a consistent proportion of the study population in the two years (Fisher exact p = 0.61) and in both seasons (Fisher exact p = 0.24). *Borrelia* infections occurred across all ages (t-test compared to all other febrile p = 0.45) (Fig. 2B) and at similar rates in male and female patients (Fisher exact p = 0.87), while male patients were more likely to test positive for malaria (Fisher exact p = 1.6e-7).

A high proportion of *Borrelia* positive patients reported generalized symptoms that were common across all febrile patients, including headache (100%), body aches (59%), and dizziness (46%), as reported in previous studies of RF *Borrelia* (Fig. 3A)^36^. *Borrelia* infection was distinguished by vomiting (49%), vomiting was observed more often in *Borrelia*-positive patients (stat = 2.8, p = 0.0026, Fisher Exact two-sided). *Borrelia*-positive patients were less likely than other febrile patients to report sore throat (7.7%, stat = 0.21, p = 0.0039, Fisher Exact two-sided). Notably, RF *Borrelia* can invade the central nervous system in severe cases; while we did not observe neurological symptoms such as seizures or loss of consciousness, one *Borrelia* positive patient reported eye pain, a potential symptom of neuroinvasive infection not observed in any other febrile patients^36,38^. A high proportion of *Borrelia*-positive patients reported contact with a febrile person (38%), contact with rats (57%), and prior travel (32%), but these exposures were common across malaria and other febrile patients and were not associated with *Borrelia* infection in particular (Fig. 3B).

**Figure 3:**
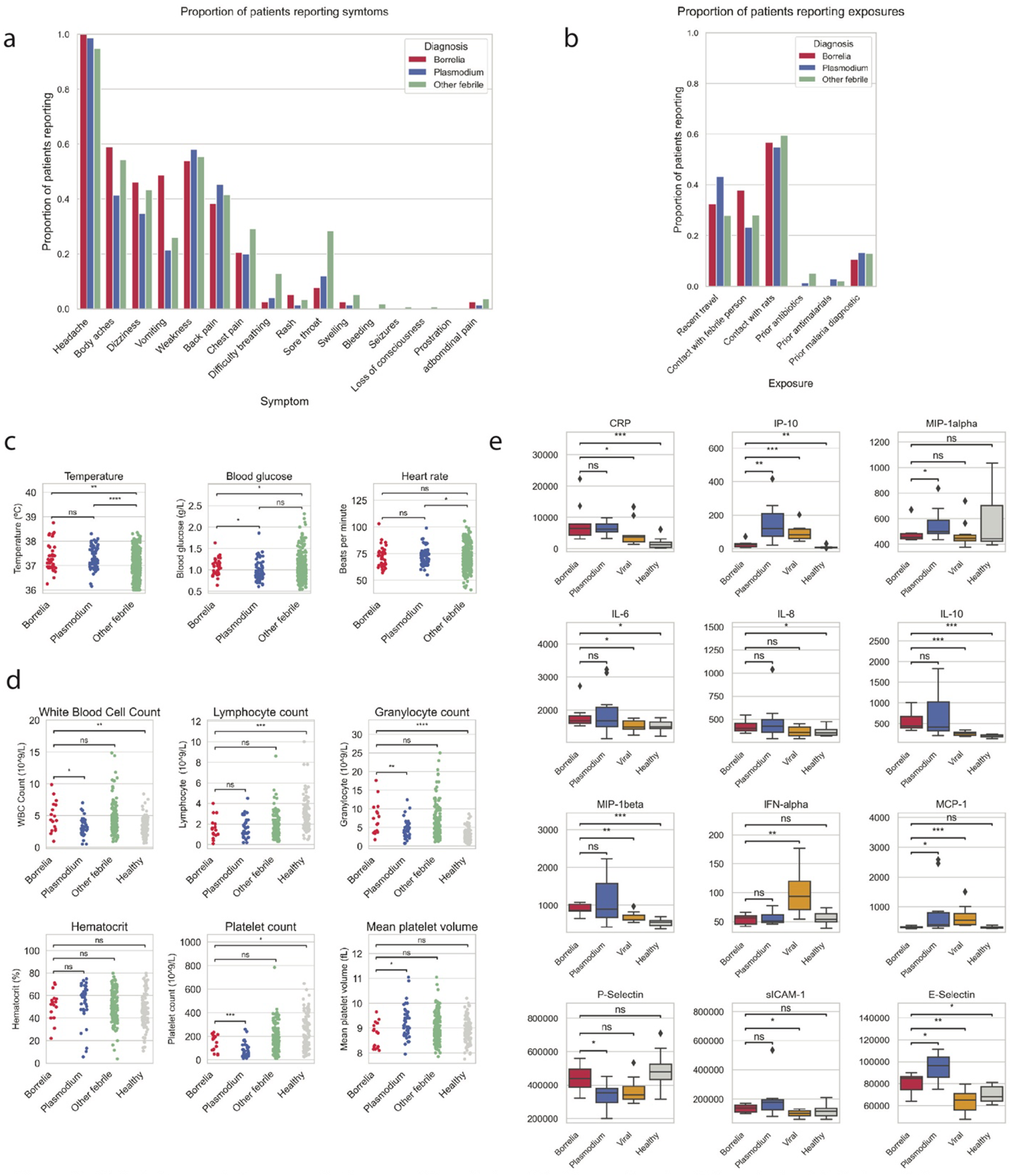
Clinical characteristics across both years of *Borrelia* (n = 39), *Plasmodium* (n = 75), and other febrile (n = 412) patients including **a.** symptoms reported by patients, **b.** exposures reported by patients, and c. vital signs. For 2019 only, **d.** blood cell counts in *Borre/ia* (n = 17), *Plasmodium* (n = 41), other febrile (n = 146), and healthy (n = 104) **e.** Serological profile of a subset of patients with *Borre/ia* (n = 9), *Plasmodium* (n = 11), viral infection (n = 10) compared to healthy patients (n = 10); cytokines/chemokines with at least one significant difference between groups shown [see Supplementary File 1 for results for all cytomkines/chemokines tested]. *Mann-Whitney-Wilcoxon test two-sided, p-value annotation legend: ns: p <= 1.00e+00, *: 1.00e-02 < p <= 5.00e-02, **: 1.00e-03 < p <= 1.00e-02, ***: 1.00e-04 < p <= 1.00e-03, ****: p <= 1.00e-04

*Borrelia* patients showed significant differences in temperature and blood glucose. On average, *Borrelia* positive patients had a higher fever (mean = 38.6°C, SD = 1.1°C) than other febrile patients (mean = 38.0°C, SD = 1.0°C, Mann-Whiteny two-sided p = 0.0011). *Borrelia* patients (mean = 1.07 g/L, sd = 0.19) were also hypoglycemic compared to other febrile patients (Other febrile: mean = 1.22 g/L, sd = 3.97, Mann-Whiteny two-sided p = 1.204e-02) (Fig. 3C).

We assessed whether the addition of a complete blood cell count with differential could help distinguish RF *Borrelia* from other febrile diseases. *Borrelia* positive patients exhibited abnormal blood counts, including lymphopenia (mean 1.5 x 10∧9 cells/L, std 1.04), granulocytosis (mean 7.47 x 10∧9 cells/L, std 4.23), and thrombocytopenia (148 x 10∧9 cells/L std 66) (Fig. 3D). However, this hematologic response was not significantly different from the response observed in other febrile patients. On a subset of patients, we measured 23 cytokines and chemokines to further distinguish *Borrelia* from other infections (Fig. 3E). *Borrelia* positive patients were distinct from viral infections in elevated CRP (Mann-Whitney two-sided, p = 0.030), IL-10 (p = 0.00038), and MIP-1Beta (p = 0.0049) and decreased IP-10 (p = 0.00070), IFN-alpha (p = 0.0022), and MCP-1 (p = 0.00038) (Fig. 3E).

Given the highly overlapping presentation of disease, we developed a weighted logistic regression model to distinguish *Borrelia* infection from other NMFI based on clinical symptoms, vital signs, and key demographic information. We evaluated the performance of the model with bootstrapping and found the model was able to predict *Borrelia* infection with high performance (recall: 0.861, 95% CI [0.837 - 0.910], precision: 0.792, 95% CI [0.696 - 0.892], and F1-score: 0.823, 95% CI [0.766 - 0.900]) (Fig. 4A). The model identified features that were significant in univariate analysis, including vomiting, temperature, blood glucose, and sore throat. Interestingly, common symptoms that were observed across all febrile patients, such as headache, body ache, and dizziness were not useful predictors on their own but were associated with increased risk of *Borrelia* when evaluated in the context of other symptoms and exposures. To test whether additional laboratory tests might further increase performance of the model, we incorporated complete blood count with differential (CBC) values. The addition of CBC values moderately increased model performance (Supplementary Fig. 9), but the difference was not significant and the test dataset was small (n = 163). When blood biomarkers were incorporated, decreased platelet count was an important predictor of *Borrelia*.

**Figure 4:**
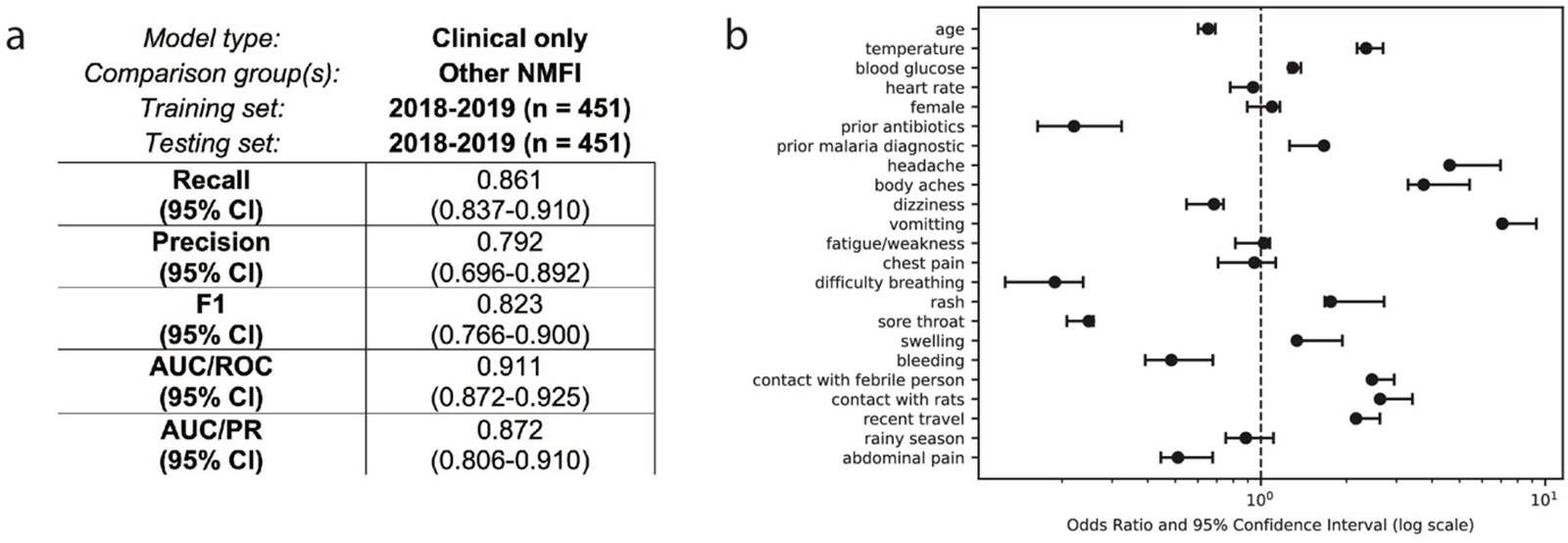
Results of best performing weighted logisistic regression model to distinguish *Borrelia* infection from other NMFI using clinical data, including demographics, symptoms, exposures, and vital signs (trained on 2018-2019 data, tested on the same data with bootstrapping and cross-validation, n = 451). a. Model performance metrics and b. odds ratios for all features retained in the final model after feature selection.

## Discussion

In this study, we employed untargeted RNA-mNGS and targeted 16S sequencing to understand the causes of acute febrile illness at an ambulatory clinic in Thiès, Senegal. As the first unbiased investigation of causes of fever in Senegal, our findings highlight the importance of looking across many pathogen types simultaneously to understand their contributions—both individually and as co-infections—to disease, and enable comparison of the strengths of genomic tools as well as clinical and epidemiological data to support disease characterization.

Arthropod-borne bacterial pathogens, *Borrelia* and *Rickettsia*, are the major identifiable causes of NMFI, but remain underdiagnosed. The frequency of these pathogens is broadly consistent with previous findings in targeted studies from other regions of Senegal^8,39^. *Borrelia* spp. associated with human and zoonotic infection have been identified in ticks across West Africa^28,36,40,41^ but human surveillance has not been done.

Our unbiased approaches and broad sampling illuminated co-infections and missed diagnoses that may be clinically relevant. Both *Borrelia* and *Rickettsia* occurred frequently as co-infections in our study. *Rickettsia* was detected more often as a co-infection with *Plasmodium* (n = 3) or *Borrelia* (n = 2) than as a stand-alone infection (n = 3). *Borrelia/Plasmodium* co-infections have been previously detected in Senegal^42^ and *in vitro* evidence suggests co-infection increases the risk of severe malarial illness^43^. *Plasmodium/R. felis* co-infections have been previously identified in Senegal^44^ and *Anopheles gambaie*, the primary malaria vector in Senegal, may be able to transmit *Rickettsia*^45^. Whether *Rickettsia* is contributing to pathogenesis or simply co-transmitting in these cases remains an open question. More research is needed to understand the interaction between the parasite and *Rickettsia* species. Given the common occurrence of co-infections and potential for negative outcomes without proper treatment of both pathogens, it is important that both surveillance and diagnostic approaches do not stop at detection of the first pathogen.

Despite using multiple methods and searching across kingdoms, we did not find any pathogen in over 70% of NMFI cases in this study. Although our enrollment criteria focused on suspicion of malaria, many of these patients reported sore throat and difficulty breathing (2019: 35%, 40/113) or abdominal pain (4.4%, 5/113). Given that only plasma samples were collected and sequenced, our approach may have missed infections limited to other body compartments, including the respiratory tract and digestive tract. Our results also demonstrate the difficulty of using RNA-mNGS to detect DNA pathogens and pathogens with different cellular compositions. Several of the bacterial infections in this cohort, particularly those with low abundance in the blood, were detected by 16S but missed by RNA-mNGS. Similarly, we were not able to resolve putative fungal hits to a suitable level of taxonomic resolution; to capture fungal pathogens, extraction methods should be optimized for lysis of fungal cells and more targeted methods such as ITS sequencing could be used. We observed wide variations in the number of *Plasmodium* reads among RDT positive patients and weak correlation with parasite density, suggesting that factors beyond parasite burden affect the number of reads recovered. There are also still many challenges in interpreting the RNA-mNGS data. In particular, RNA-mNGS is highly susceptible to contamination, such as the novel *Reoviridae* RdRp sequences we detected across febrile cases, healthy controls and non-template controls. Negative controls should be included alongside samples throughout processing to measure the “contaminome,” which likely differs between sample types and labs^15^.

We find that combinations of clinical signs and symptoms can increase suspicion for *Borrelia* and support targeting clinical care. Due to the lack of available point-of-care tests, many clinics in LMICs rely on malaria RDTs as the primary diagnostic for febrile illness and may give blanket antibiotic treatment to RDT-negative patients^46^. Concerns have been raised about this practice^47^, including driving antibiotic resistance and exposing patients to unnecessary risks or side effects. For example, tetracycline antibiotic treatment—the recommended therapeutic for *Borrelia* and *Rickettsial* infections—poses the risk of Jarich-Herxheimer reaction (JHR), a severe inflammatory response to spirochete lysis that often requires close patient monitoring^48^. We identify a number of clinical and immunological features that could bolster support for a *Borrelia* diagnosis. Our weighted logistic regression model indicates that when larger cohorts and diverse data types are aggregated, *Borrelia* can be differentiated from other NMFI with reasonable accuracy. The addition of laboratory measures, such as CBC, did not substantially improve differential diagnosis. Further, data from a subset of our cohort supports the hypothesis that a chemokine panel including TRAIL, IP-10 and CRP, previously employed for detection of bacterial infections in hospitalized children^49^, could apply more broadly and help distinguish bacterial infections such as *Borrelia* from other causes of febrile illness in the ambulatory setting. However, none of these approaches are currently actionable as point-of-care tests and their lower precision would still necessitate confirmatory diagnostic testing.

The challenge of quickly and accurately differentiating *Borrelia* infections underscores the need for improved diagnostics. The diagnostic gold standard for *Borrelia* remains microscopic examination of blood smears, which has low sensitivity in our study and others^35^. Further, smear cannot distinguish species, and is highly dependent on a trained practitioner. Our study showed that molecular methods, including 16S sequencing and mNGS, can detect *Borrelia* infections, but are not practical in the clinical setting. Increased availability of qPCR assays and improved point of care nucleic acid diagnostics could enable detection and treatment of *Borrelia* across differently resourced clinical settings and improve our understanding of the geographic range and diversity of this pathogens

An increasingly heterogeneous pathogen landscape will require the integration of targeted diagnostics and broad surveillance to effectively combat acute febrile infections moving forward. Decreased malaria transmission coupled with changes in the range and incidence of other pathogens—influenced in part by global climatic shifts—is broadening the array of pathogens accounting for the majority of febrile disease^50^. Fully understanding this landscape will require surveillance systems utilizing unbiased approaches that have been validated for viral, bacterial and eukaryotic pathogens and diverse sample types. In tandem, for *Borrelia* and other pathogens known to frequently cause NMFI, the development and availability of diagnostic tests that are cheap, rapid and sensitive will be key to enable appropriate clinical treatment at the point of care and support deeper investigations of the pathophysiology of disease.

## Methods

### Sample collection

Febrile cases were selected from patients presenting to the SLAP outpatient clinic in Thies, Senegal during the collection period under local IRB (SEN15/46) and Harvard IRB (IRB19-0023). Informed consent was obtained for all enrollees; for minors under 18 years of age or individuals unable to provide their own consent, the consent of a parent or legal guardian was obtained. The study team explained to potential participants that their participation was strictly voluntary and that they could withdraw from the study at any time without any penalties or consequences and translations were made for potential participants (or their parent / legal guardian) that do not understand or cannot read the language in which the consent form was produced.

Patients who met the following inclusion criteria and gave consent were enrolled: (1) Febrile symptoms within the 3 days up to and including the day of presentation, (2) Age 2-75 years, and (3) Ambulatory with no signs of severe malarial (glucose < 2.2mM, hemoglobin < 5 gms/dL). A subset of DENV positive samples from 2018 were previously sequenced and published^4^ and were excluded from this study other than being used as a viral outgroup in the chemokine/cytokine analyses. At the time of enrollment, a structured interview including personal information, demographics, and self-reported symptoms was completed, vital signs were measured and blood was drawn by a trained practitioner. *Plasmodium falciparum* RDT (Bioline Malaria Ag P.f., Abbott), thick blood smear, and thin blood smear was performed and blood glucose (HemoCue 201/301) and hemoglobin (HemoCue Glucose 201) were measured.

### Sample processing

Blood samples were stored on ice upon collection (maximum 8 hours). In the lab, blood samples were split into two aliquots. 200 ul of the first aliquot was used for an automated blood count with differential (Mindray BC 20s) and 200uL was used for a blood spot (Whatman filter paper) The remaining aliquot was centrifuged at 2300 rpm for 10 min to separate plasma, buffy coat and red blood cells. 140 uL of plasma was transferred to a tube containing 760uL of inactivation buffer (AVL, Qiagen). The remaining plasma was stored without inactivation. Red blood cells, buffy coat, plasma, and inactivated plasma were flash frozen and stored at -80°C until further processing. Total nucleic acids were extracted with Qiagen QIAmp Viral Mini RNA according to the manufacturer’s protocol. Total nucleic acids were split into two aliquots and treated with Lucigen RNAse I or Ambion TURBO DNase to obtain purified DNA and RNA, respectively.

### Unbiased RNA sequencing

#### Library preparation and sequencing

Unbiased RNA sequencing libraries were prepared as described previously^51^. Briefly, cDNA synthesis was performed from DNAse-treated RNA using random hexamers (Illumina) and SSIV. Libraries were prepared from cDNA using Nextera XT (Illumina) and UD index primers (IDT/Illumina) with the following modifications to account for low cDNA input: the volume of ATM was decreased from 1uL to 0.5uL per 10uL reaction and PCR cycles were increased to 17 cycles. Libraries were purified using AMPure XP (Agilent), quantified using KAPA Biosystems Universal Library Quantification, and pooled equally for 75bp paired end sequencing on NovaSeq SP to obtain at least 2 million reads per sample.

#### Taxonomic classification of RNA-mNGS reads

Unbiased RNA sequencing reads were processed using viral-ngs^52^. Briefly, reads were demultiplexed, adapter sequences were trimmed, and sliding-window quality filtering was performed. Human reads were filtered out using BMTAGGER and BLASTN. Cleaned reads were uploaded to NCBI SRA under BioProject PRJNA662334 (Accession numbers: SRR24622550-SRR24622641, SRR24995052-SRR24995258). Samples with insufficient reads (<2 million) were removed from further analysis (n = 1, SHC1064, Accession: SRR24995248).

Taxonomic classification of human-depleted reads was performed using Kraken2^53^ with the JHU PlusPF database (JHU, downloaded 12-13-2022). Kraken2 results were thresholded to consider only viral genera with at least 5 reads/million raw reads. Results were filtered to remove viral taxa detected in non-template controls (at least one non-template control) and healthy patients (at least 2 healthy patients). Further, viral genera with non-vertebrate hosts were filtered out.

To verify Kraken2 classifications, we performed a protein-sequence similarity search using DIAMOND-blastx^54^. Cleaned, de-duplicated reads were used for *de novo* contig assembly with SPAdes and DIAMOND-blastx was run on all *de novo* contigs > 100 bp long with the complete nr database (downloaded December 2022). The least common ancestor of the top e-value hits was identified using a custom script (lakras/bio-helper-scripts/blast/retrieve_top_blast_hits_LCA_for_each_sequence.pl). A classification was considered verified by DIAMOND if the cumulative length of contigs with a high identity match (mean identity of top e-value hits >= 95%) to the expected viral genus was at least 1kb. After DIAMOND-blastx verification, raw reads were aligned to the NCBI Virus RefSeq and assessed for evenness and depth of coverage.

To assess for presence of divergent viral taxa that are not well represented in the database and may be missed by Kraken2, DIAMOND-blastx results were filtered for viral hits with low identity (20-80% amino acid identity). Results were filtered for viral families with a per-sample cumulative *de novo* contig length >= 500 bp. *Narnaviridae* and *Reoviridae* sequences were aligned with all available RdRp sequences for each family in nr using MAFFT and trimmed with trimAl. Maximum likelihood phylogenetic trees were generated in IQ-TREE^55^ with a GTR-gamma substitution model and bootstrapping (n = 1000) and visualized in FigTree v1.4.4^56^.

#### Viral genome assembly and phylogenetic analysis

For virus-positive samples, reference-guided *de novo* assembly was performed using viral-ngs (assembly with SPAdes, reference-assisted improvements with MUMMER and MUSCLE/MAFFT and gap sealing with Gap2Seq). The NCBI Virus RefSeq as well as any >80% complete genomes available from Senegal were provided as references. For samples with near-complete or complete (>80%) genomes, genotype was determined with GenomeDetective (DENV, HBV) or by phylogenetic tree (Parvovirus B-19, GBV-C). Phylogenetic trees were generated with all >80% complete references from Africa downloaded from NCBI viruses (DENV1, DENV3, HBV), or all global >80% complete genomes available in NCBI virus (Parvovirus B-19, GBV-C). Sample sequences and reference sequences were aligned using MAFFT v7^57^ and alignments were trimmed with trimAl v1.4.rev15^58^ to remove any bases with coverage in <80% of sequences. Maximum likelihood phylogenetic trees were generated in IQ-TREE^55^ with a GTR-gamma substitution model and bootstrapping (n = 1000). Visualizations were generated with FigTree v1.4.4^56^.

#### Plasmodium RNA-mNGS analysis

In order to quantify *Plasmodium* rpm, the number of reads classified as *Plasmodium* at the genus level by Kraken2 was divided by the raw read count in millions. The cut-off for considering a sample positive for *Plasmodium* by mNGS was determined by calculating the 99th percentile for healthy control samples, 550 *Plasmodium* rpm. Cleaned deduplicated reads were aligned to the *P. falciparum pfhrp2* (PlasmoDB PF3D7_0831800) with NovoAlign.

#### Fungal RNA-mNGS analysis

In order to detect possible fungal infections, we calculated the rpm classified as Fungi by Kraken2^53^ at the kingdom level and determined the 99th percentile for healthy controls, 3399 rpm. We identified 5 samples (4 febrile, 1 control) with high fungal reads. We attempted taxonomic classification of these reads with Kraken2 at the genus level and saw hits across multiple genera for many samples (Supplementary Fig. 8B). To try to improve fungal classification, we performed a nucleic acid search of all *de novo* contigs from these 5 samples with megablast against nt (Supplementary Fig. 8C) and a translated nucleic acid search with DIAMOND-blastx^54^ (as described above). For both searches, we found the least common ancestor of the top hits (as described above), and filtered hits in the kingdom Fungi (taxid: 4751) with >90% identity and >30% coverage.

### qPCR

Total bacterial load was quantified from extracted plasma DNA with primers targeting the V1-2 region (see Supplementary Table 2 for primer sequences) with Power SYBR Green PCR MasterMix (Thermo Fisher) under the following conditions: 300nM each F/R primer, 95°C hold for 10 min, 40 cycles of 95°C for 15 sec, 50°C for 1 min, 75°C for 30s. Pan-*Borrelia* qPCR was performed with Power SYBR Green PCR MasterMix (Thermo Fisher) under the following conditions: 400 nM each F/R primer, 95°C hold for 10 min, 40 cycles of 95°C for 15 sec, 60°C for 1 min.

### 16S sequencing

#### Library preparation and sequencing

Samples with high bacterial load (V1-2 qPCR CT < 31.5) were selected for 16S sequencing (2019: febrile n = 129, healthy n = 35, 2018: febrile n = 84). Libraries were generated by amplification with tailed universal primers Tail-V1-2F and Tail-V1-2R (Supplementary Table 2) targeting variable regions 1-2. Amplification was performed on 5ul of RNAse-treated DNA using Q5 high fidelity polymerase (NEB) with forward and reverse primers at 100nM each with the following cycling conditions: 98°C for 30 sec; 35 cycles of 95°C for 15 sec and 63°C for 2 min, 4°C hold. PCR products were cleaned using Ampure XP (Agilent) with 0.7X ratio to remove primer dimers. Adapters and barcodes (BroadDuplex Seq) were added with a second PCR reaction using Q5 polymerase and the following cycling conditions: 98°C for 30 sec; 18 cycles of 95°C for 15 sec, 60°C for 15S, 72°C for 30S, followed by 72°C for 5min, 4°C hold. A subset of libraries were visualized with hsDNA BioAnalyzer (Agilent) and all libraries were quantified with KAPA Biosystems Universal Library Quantification Kit before pooling equally across samples and 250 bp paired-end sequencing on Illumina MiSeq v2 with 40% PhiX, given the low diversity of the single-amplicon library.

#### Analysis

Sequencing reads were demultiplexed using viral-ngs and demultiplexed FASTQ files were imported into qiime2^59^ for further analysis. Briefly, after sliding window quality filtering and adapter trimming with cut-adapt (minimum-length 20), paired reads were joined with vsearch (tuncqual 15, minlen 35, minovlen 10, maxdiffs 3), and ASVs were generated with Deblur (trim-length 280). ASVs were taxonomically classified by blastn search of ASV sequences against the NCBI 16S rRNA db (downloaded June 2023). The least common ancestor of the top e-value hits was determined using a custom script (lakras/bio-helper-scripts/blast/retrieve_top_blast_hits_LCA_for_each_sequence.pl). Abundance was calculated by dividing the number of ASVs in each genus by the total number of ASVs in each sample. Abundance data was filtered to show only taxa accounting for greater than 5% of ASVs. Taxa detected in non-template controls (n >= 1) and healthy controls (n >= 2) were filtered out.

For *Borrelia* and *Rickettsia* positive samples, sample ASVs were aligned with MAFFT^57^ (Geneious) with curated high-quality 16S sequencing from the NCBI 16S rRNA BioProject (BioProject: 33175) from each genus respectively. Alignments were trimmed with trimAl to remove positions with gaps in >90% of sequences and a maximum-likelihood phylogenetic tree was generated using IQ-TREE^55^ with bootstrapping (n = 1000) and annotated with FigTree v1.4.4^56^.

### IGS Sequencing

The intergenic spacer (IGS) region was amplified using a nested-PCR. First, the region was amplified from 5uL of extracted DNA with primers targeting the IGS region (IGS-outer F/R, see Supplementary Table 2) using Q5 polymerase and the following cycling conditions: 98°C for 30 sec; 35 cycles of 94°C for 30 sec, 66°C for 30S, 74°C for 60s, followed by 74°C for 2 min, 10°C hold. After a 0.75X AMPure XP cleanup to remove excess primer, a nested PCR was performed (IGS-inner F/R, see Supplementary Table 2) using Q5 polymerase and the following cycling conditions: 98°C for 30 sec; 30 cycles of 94°C for 30 sec, 67°C for 30S, 74°C for 60s, followed by 74°C for 2 min, 10°C hold. Amplified product was cleaned with AMPure XP (0.75X), visualized with BioAnalyzer TapeStation and bi-directional Sanger sequencing was performed (Azenta). Paired Sanger sequencing traces were analyzed in Geneious to generate a consensus sequenced and aligned with all available reference sequences for *B. crocidurae, B. duttonii, and B. recurrentis* using MAFFT^57^. A maximum-likelihood phylogenetic tree was generated using IQ-TREE^55^ and annotated with FigTree v1.4.4^56^.

### Blood smear examination

Thick and thin blood smears were fixed with methanol and stained with 3mL 10% Giemsa for 10 minutes. Stained smears were examined under a brightfield microscope for evidence of *Borrelia*. Slides were first scanned at low (40X) magnification and then at high (100X) magnification with oil immersion to quantify organisms per field (representative image, Supplementary Fig. 6B).

### Serology

We assessed levels of common cytokines and chemokines using the Luminex platform on plasma from a subset of patients and controls. Specifically, we selected 10 healthy controls and 30 cases representing 10 each with a confirmed diagnosis of; malaria (based on malaria RDT administered at enrollment), Borrelia (based on pan-*Borrelia* qPCR^37^), or viral infection. Samples with a viral infection were all confirmed to have Dengue virus and are a subset of those cases previously described elsewhere^4^. Cases were selected randomly within those meeting each criteria. All samples were collected in 2018. We used the Inflammation 20-Plex Human ProcartaPlex™ Panel (Invitrogen), according to the manufacturer’s instructions. We additionally performed a 2-plex assay for Human IP-10 and TRAIL (using IP-10 and TRAIL ProcartaPlex™ simplex kits from Invitrogen); two markers previously found to differentiate bacterial infections in hospitalized children^49^, as these were not included on the predefined panel. All samples were run in duplicate on a Luminex MAGPIX® instrument in Senegal. We confirmed that standards showed the expected values and calculated the average across duplicates, which we report in the text.

### Multivariate model

We developed a weighted logistic regression model that considered 46 candidate predictors based on clinical symptoms, vital signs, and key demographic information to predict *Borrelia* infection. Before modeling, the dataset underwent the following preprocessing steps: imputation of missing values with the Multivariate Imputation by Chained Equations (MICE) algorithm from the fancyimpute Python library^60^, one hot-encoding of categorical features, and standardization of continuous features using the StandardScaler from the scikit-learn library. Given that normal heart rate varies significantly with age, heart rates were binned and treated as a categorical variable. Heart rate categories were high, low, or normal based on the Pediatric Advanced Life Support guidelines as follows: for age 3-5, normal range 80-120, age 6-11 normal range 75-118, age 12+ normal range 60-100^61^.

The sample size was relatively small, with 526 patients and only 38 q-PCR confirmed cases of *Borrelia* infection. Given this inherent class imbalance in the dataset, a combination of techniques was employed to address this challenge. The Synthetic Minority Oversampling Technique (SMOTE) was applied to generate synthetic samples for the minority class (q-PCR confirmed *Borrelia* case)^62^. This technique mitigates class imbalance by interpolating between existing samples and creating synthetic instances of the minority class. During model training, class weights were also applied to adjust the influence of each class. The class weights were inversely proportional to the class frequencies, with higher weights assigned to the minority class.

A logistic regression model was chosen for its interpretability and ability to estimate the probabilities of class membership. After a univariate screening for statistically significant predictors using chi-squared test (p-value < 0.05), several Feature Selection techniques were applied to construct the final set of predictors in the model. Before training the model, feature importance was assessed using a combination of three different feature selection methods; mutual information classification^63^, Recursive Feature Elimination^64^, and Lasso L1 regularization^65,66^ to ensure consistent elimination of redundant variables. The selected features were used in the final weighted logistic regression model to analyze the performance of *Borrelia* prediction. The regularization parameter from Lasso was tuned using cross-validation of different alpha values and the best alpha value was used to train the final model and evaluate its performance on the test set.

Model performance was evaluated with bootstrapping and 5-fold cross validation to quantify optimism and generalizability. The optimism-corrected metrics include F1-score, precision, recall, area under the ROC curve (AUC-ROC), and area under the precision-recall curve (AUC-PR). Odds ratios and their corresponding confidence intervals for all the model coefficients were calculated to assess the impact of predictor variables on the odds of the outcome. All predictive analyses were conducted using the Python programming language. The scikit-learn library was utilized for preprocessing, model training, and evaluation. Statistical analyses and visualization of odds ratios and confidence intervals were done using the R statistical package.

## Data Availability

RNA-mNGS short read sequence data have been deposited in NCBI SRA and Genbank databases under BioProject PRJNA662334 Accession numbers SRR24622550-SRR24622641, SRR24995052-24995258. 16S v1-2 amplicon sequencing data and assembled viral genomes will be deposited shortly under BioProject PRJNA662334, data currently available upon request. The Python source code for the multivariate Borrelia prediction model is available at github.com/colabobio/borrelia-diagnosis-prediction-models.

## Author contributions

According to the CRediT format, author contributions are as follows: Conceptualization: A.B.D., A.S.B., Z.C.L., N.S., P.C.S., K.J.S., and D.N.; Methodology: A.B.D., A.S.B., Z.C.L., N.S., P.C.S., K.J.S., and D.N.; Software: D.J.P. ; Validation: P.C.S., K.J.S., and D.N.; Formal Analysis: Z.C.L, K.J.S., W.M., and A.C.; Investigation: Z.C.L, A.S., A.G., T.N., M.S., Y.D., A.M., and I.N.; Data Curation: J.G., M.Ndiop, D.S.; Writing – Original Draft: Z.C.L. Writing - Review & Editing: K.J.S., S.F.S., B.M, and P.C.S. with input from all authors; Visualization: Z.C.L., W.M.; Supervision: P.C.S., K.J.S., and D.N.; Project Administration: M.F.P., M.Ndiaye.; Funding acquisition: P.C.S., D.N., K.J.S., D.J.P.

## Data and Code Availability

RNA-mNGS short read sequence data have been deposited in NCBI’s SRA and Genbank databases under BioProject PRJNA662334 Accession numbers SRR24622550-SRR24622641, SRR24995052-24995258. 16S v1-2 amplicon sequencing data and assembled viral genomes will be deposited shortly under BioProject PRJNA662334, data currently available upon request. The Python source code for the multivariate Borrelia prediction model is available at github.com/colabobio/borrelia-diagnosis-prediction-models.

## Acknowledgements

We thank the Programme National de Lutte contre le Paludisme (National Malaria Control Program) for providing malaria surveillance data, and Fama and Astou for support with data entry and organization. We thank Lydia Krasilnikova for advice and custom scripts for BLAST analysis, and Flavia Negrete for support with data uploads and sharing. We thank Jacob Lemieux and Gordon Adams for providing valuable guidance on *Borrelia* sequencing.

