## Supplementary Figures for "Improving diagnosis of non-malarial fevers in Senegal: *Borrelia* and the contribution of tick-borne bacteria"

### Supplementary Material:

**Supplementary Table 1: Viral pathogens detected in 2019**

| Virus | Genotype | Season | Patient age (years) |
|---|---|---|---|
| Dengue Virus 1 | Related to but not part of Genotype III | Dry Season | 22 |
| Dengue Virus 3 | Genotype III | Wet Season | 29 |
| Hepatitis B Virus | Genotype A | Dry Season | 10 |
| Hepatitis B Virus | Genotype E | Wet Season | 12 |
| HIV-1 | | Dry Season | 24 |
| Pegivirus C | Genotype I | Dry Season | 48 |
| Pegivirus C | Genotype I | Wet Season | 10 |
| Pegivirus C | Genotype I | Wet Season | 33 |
| Pegivirus C | Genotype I | Wet Season | 26 |

**Supplementary table 2: Primer sequences**

| ***Name*** | ***Sequence*** | ***Assay*** | ***Reference*** |
|---|---|---|---|
| *V1/2-F* | 5'-CTGCTGCCTCCCGTAGGAGT-3' | Total bacterial load qPCR | Kingry, 2020[19] |
| *V1/2-R* | 5'-AGAGTTTGATCCTGGCTCAG-3' | | |
| *Borrelia-F* | 5'-AGCYTTTAAAGCTTCGCTTGTAG-3' | pan-Borrelia qPCR | Kingry, 2018[37] |
| *Borrelia-R* | 5'-GCCTCCCGTAGGAGTCTG-3' | | |
| *Tail-V1/2-F* | 5'-ACACTCTTTCCCTACACGACGCTCTT CCGATCTCTGCTGCCTCCCGTAGGAGT-3' | 16S sequencing library construction | |
| *Tail-V1/2-R* | 5'-GTGACTGGAGTTCAGACGTGTGCTCTT CCGATCTAGAGTTTGATCCTGGCTCAG-3' | | |
| *IGS-outer-F* | 5'-GTATGTTTAGTGAGGGGGGTG-3' | IGS sequencing | Bunikis, 2004[31] |
| *IGS-outer-R* | 5'-GGATCATAGCTCAGGTGGTGAG-3' | | |
| *IGS-inner-F* | 5'-AGGGGGGTGAAGTCGTAACAAG-3' | | |
| *IGS-inner-R* | 5'-GTCTGATAAACCTGAGGTCGGA-3' | | |

a

| | | 2018 | | | | 2019 | | | |
|---|---|---|---|---|---|---|---|---|---|
| | | ***Febrile*** | | ***Healthy*** | | ***Febrile*** | | ***Healthy*** | |
| *Female* | *Adult (18+)* | 79 | (24.6%) | 151 | (38.8%) | 51 | (25.0%) | 32 | (30.8%) |
| | *Adolescent (13-17)* | 26 | (8.1%) | 31 | (8.0%) | 14 | (6.9%) | 6 | (5.8%) |
| | *Child (6-12)* | 26 | (8.1%) | 42 | (10.8%) | 20 | (9.8%) | 13 | (12.5%) |
| | *Young child (2-6)* | 8 | (2.5%) | 15 | (3.9%) | 8 | (3.9%) | 6 | (5.8%) |
| *Male* | *Adult (18+)* | 97 | (30.2%) | 45 | (11.6%) | 59 | (28.9%) | 21 | (20.2%) |
| | *Adolescent (13-17)* | 22 | (6.9%) | 33 | (8.5%) | 14 | (6.9%) | 10 | (9.6%) |
| | *Child (6-12)* | 42 | (13.1%) | 48 | (12.3%) | 31 | (15.2%) | 8 | (7.7%) |
| | *Young child (2-6)* | 21 | (6.5%) | 24 | (6.2%) | 7 | (3.4%) | 8 | (7.7%) |
| | ***Total*** | **321** | | **389** | | **204** | | **104** | |

b

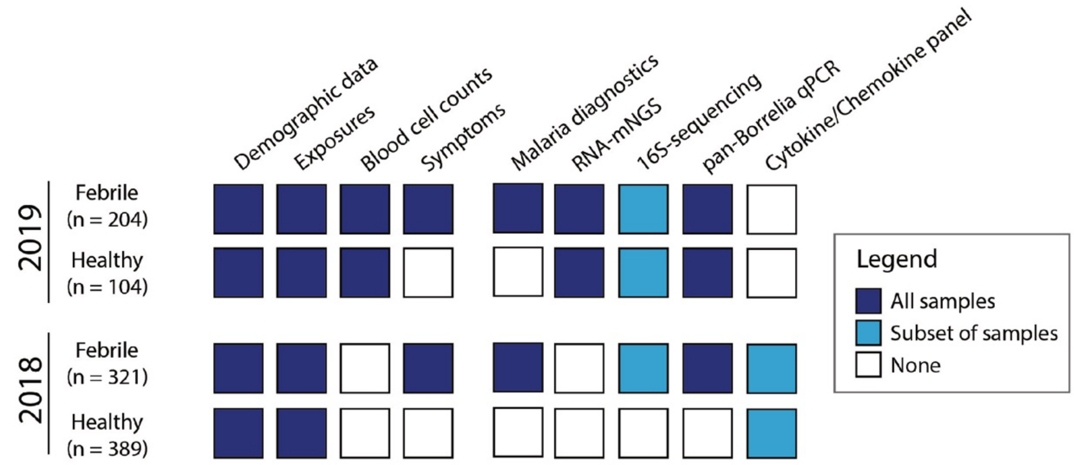


**Supplementary Figure 1:** Overview of data set in the study **a.** Demographics of febrile patients and healthy controls enrolled in the study from 2018-2019. **b.** Summary of metadata, clinical diagnostics, and lab data available for febrile cases and healthy controls in 2018 and 2019.

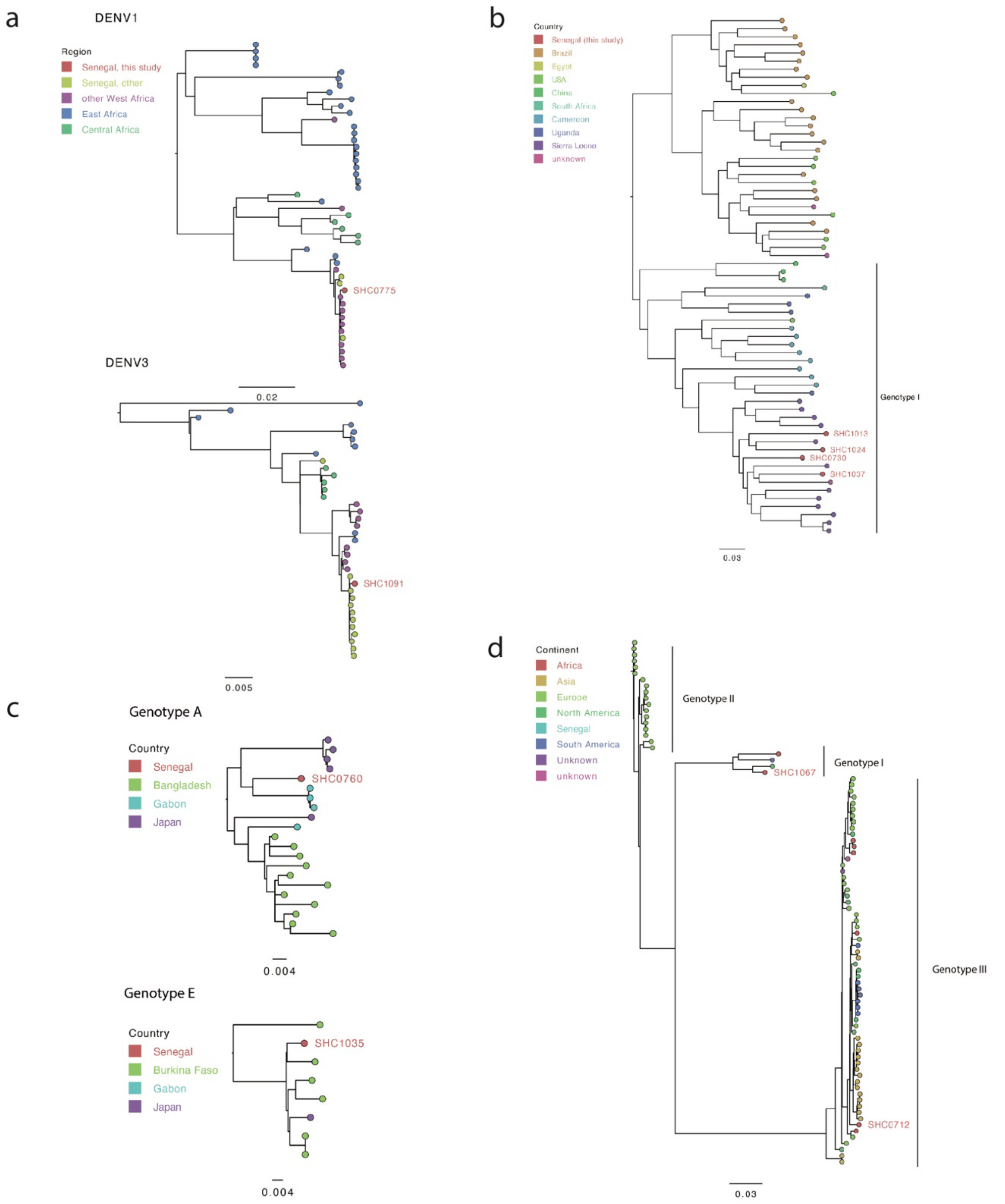


**Supplementary Figure 2:** Viral isolate phylogenetic trees **a.** Maximum likelihood phylogenetic tree (IQ-TREE) of DENV1 and DENV3 isolates from this study (red) in the context of all > 80% complete genomes from Africa in NCBI virus. **b.** Maximum likelihood phylogenetic tree (IQ-TREE) of all >80% complete Pegivirus C genomes colored by country of origin. **c.** Maximum likelihood phylogenetic tree (IQ-TREE) of Hepatitis B isolates from this study (red) in the context of all >80% complete Genotype A or Genotype E Hepatitis B virus genomes from Africa in NCBI virus. **d.** Maximum likelihood phylogenetic tree (IQ-TREE) for Parvovirus B19 isolates from this study (red) in the context of all >80% complete Parvovirus B19 genomes from a human host in NCBI virus.

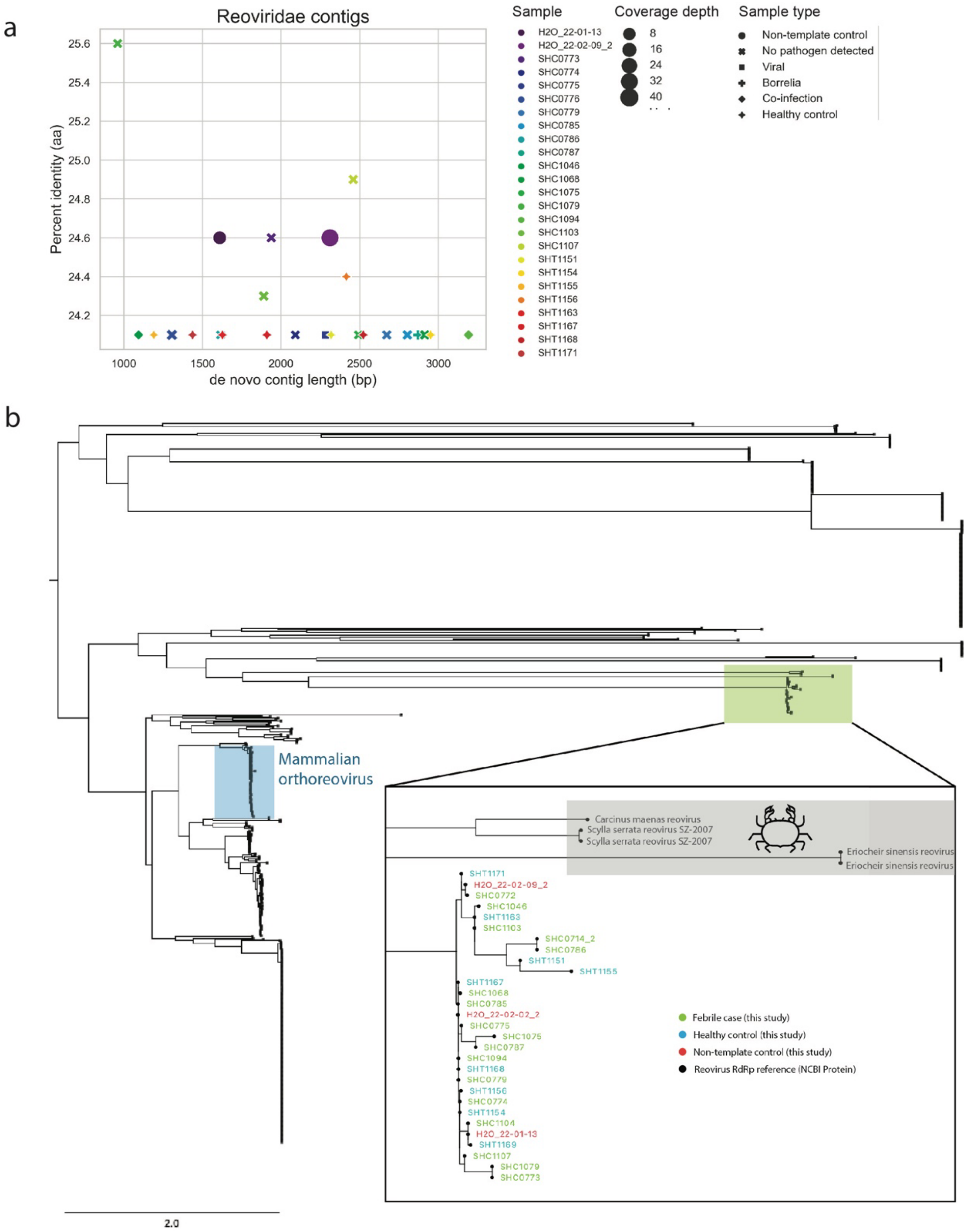


**Supplementary Figure 3:** Divergent *Reoviridae de novo* contigs identified in RNA-mNGS with **a**. percent amino acid identity to the closes DIAMOND-blastx match on the y-axis and contig length on the x-axis. Each dot represents a contig, colored by the sample ID with shape indicating the type of sample and size of the marker indicating the mean read depth across the contig. **b.** Maximum likelihood phylogenetic tree of available RdRp amino acid sequences for *Reoviridae* and *de novo* contigs from febrile patients (green), healthy controls (blue) and non-template controls (red) from this cohort.

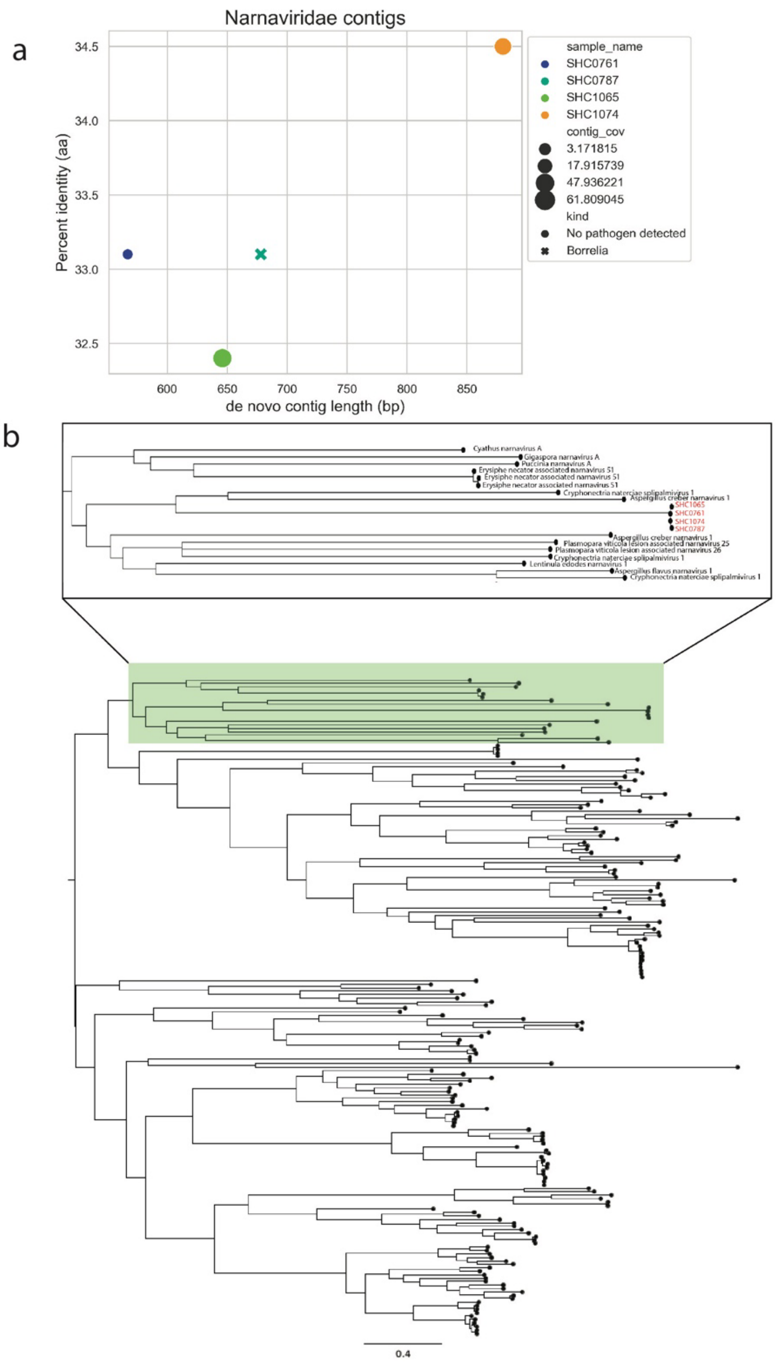


**Supplementary Figure 4:** Divergent *Narnaviridae de novo* contigs identified in this cohort with **a.** percent amino acid identity to the closes DIAMOND-blastx match on the y-axis and contig length on the x-axis. Each dot represents a contig, colored by the sample ID with shape indicating sample type and the size of the marker indicating the mean read depth across the contig. **b.** Maximum likelihood phylogenetic tree of available RdRp amino acid sequences for *Narnaviridae* and *de novo* contigs from febrile patients in this cohort (red).

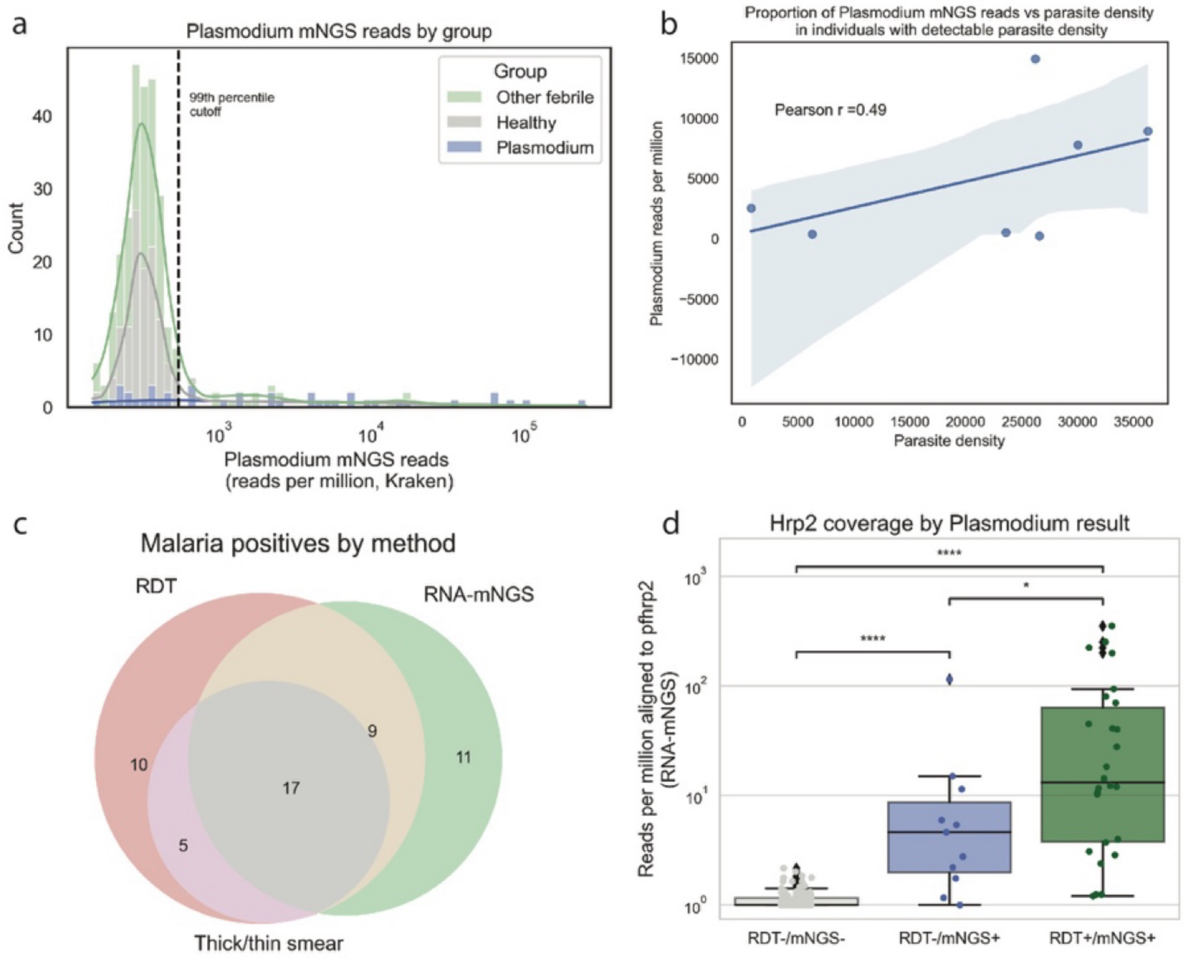


**Supplementary Figure 5: a.** Distribution of RNA-mNGS read proportion in healthy, *Plasmodium* RDT(+) febrile and *Plasmodium* RDT(-) febrile patients with 99th percentile of *Plasmodium* abundance in healthy individuals, the threshold for considering sample *Plasmodium* positive by RNA-mNGS, marked by dashed line. **b.** *Plasmodium* RNA-mNGS reads per million raw reads vs parasite density for thick/thin blood smear positive patients with detectable parasitemia. **c.** *Plasmodium* detection by rapid diagnostic test (RDT), RNA-mNGS, or thick/thin smear. **d.** Reads per million raw reads aligned to *pfhrp2* (PlasmoDB PF3D7_0831800) , the gene encoding HRP-2, the target antigen target for the *P. falciparum* RDTs.

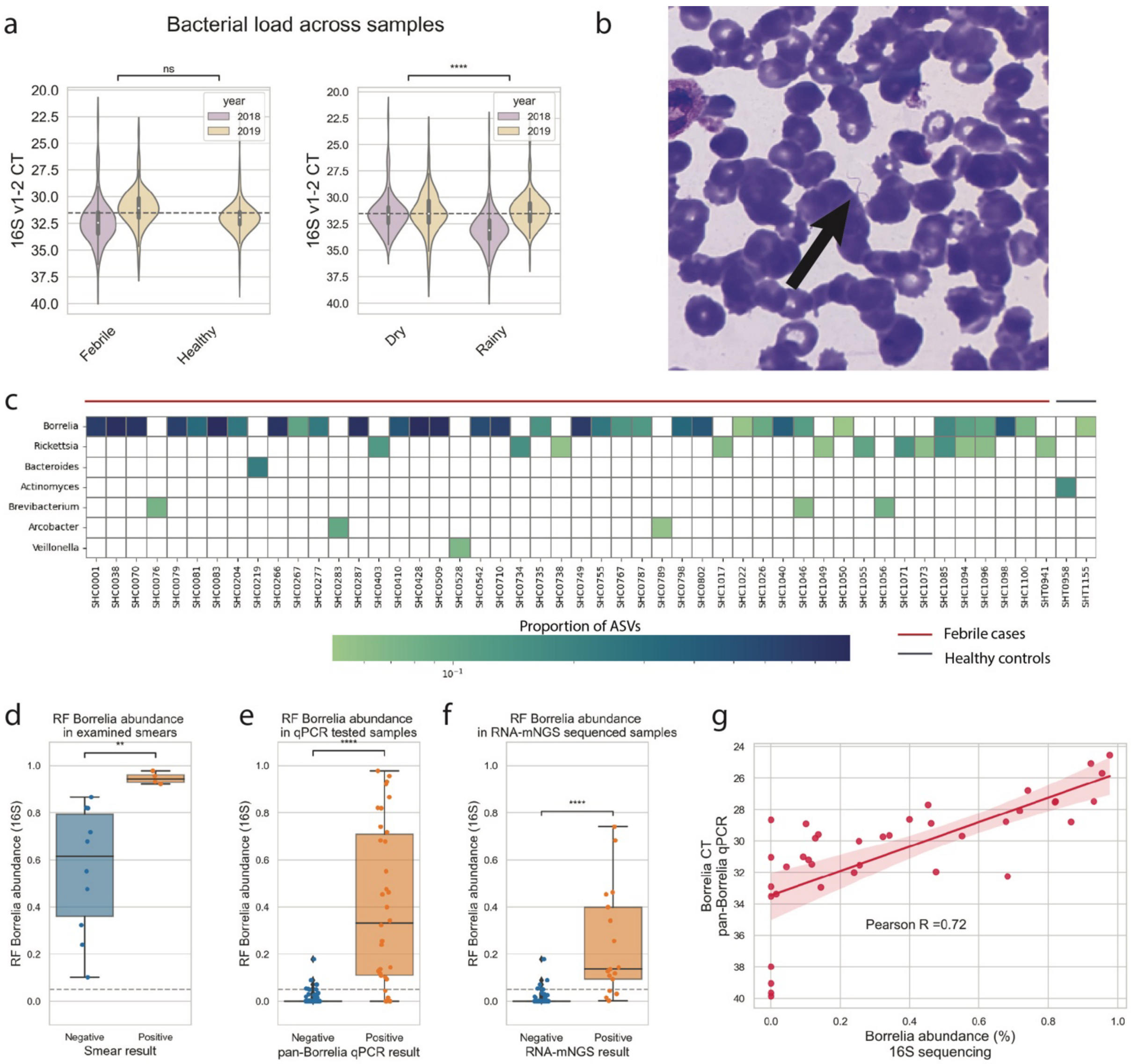


**Supplementary Figure 6: a.** Bacterial load across samples measured by eubacterial V1-2 qPCR; dashed line indicates the 31.5 CT cutoff used to select samples for 16S sequencing. **b.** Representative Giemsa stained blood smear (100X magnification) from a *Borrelia* smear positive patient with arrow indicating the spirochete. **c.** Bacterial pathogens identified by 16S across both years in febrile cases (red line) and healthy controls (grey line). *Borrelia* abundance by 16S (ASVs classified as *Borrelia* at the genus level / total ASVs in sample) for samples positive and negative for *Borrelia* by **d.** blood smear examination, and **e.** RNA-mNGS, and **f.** pan-*Borrelia* qPCR **g.** Quantification of *Borrelia* load by pan-*Borrelia* qPCR (y-axis) vs v1-2 1 ASV abundance (x-axis) *Mann-Whitney-Wilcoxon test two-sided, p-value annotation legend: ns: p <= 1.00e+00, *: 1.00e-02 < p <= 5.00e-02, **: 1.00e-03 < p <= 1.00e-02, ***: 1.00e-04 < p <= 1.00e-03, ****: p <= 1.00e-04

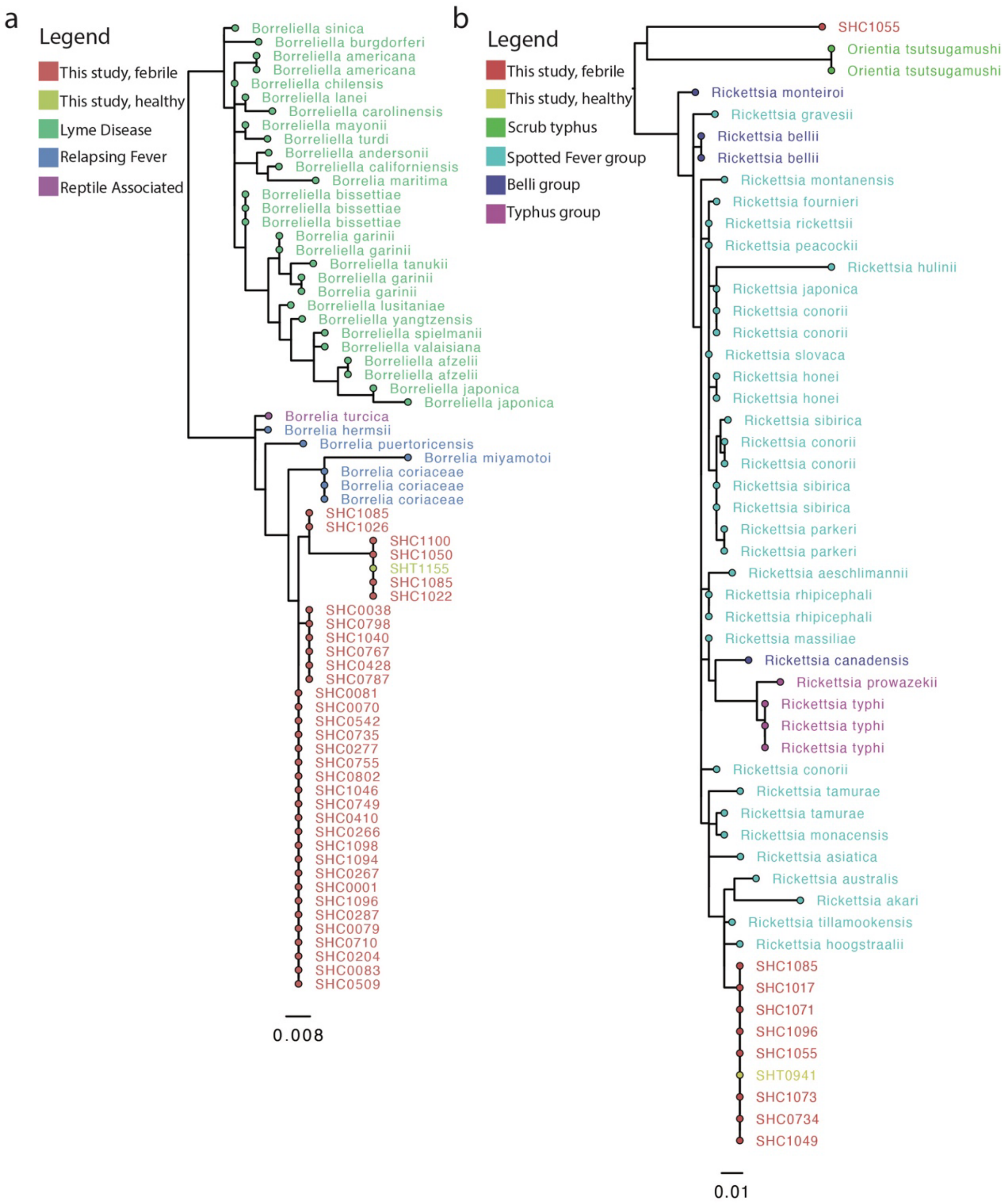


**Supplementary Figure 7:** Midpoint rooted maximum likelihood phylogenetic trees (IQ-TREE) of 16S v1-2 rRNA gene v1-2 sequences for **a.** *Borrelia* isolates from this study (febrile: red, healthy: yellow) in the context of *Borreliella* and *Borrelia* sequences from the curated NCBI 16S rRNA target loci project and **b.** *Rickettsia* isolates from this study (febrile: red, healthy: yellow) in the context of *Rickettsia* sequences from the curated NCBI 16S rRNA target loci project.

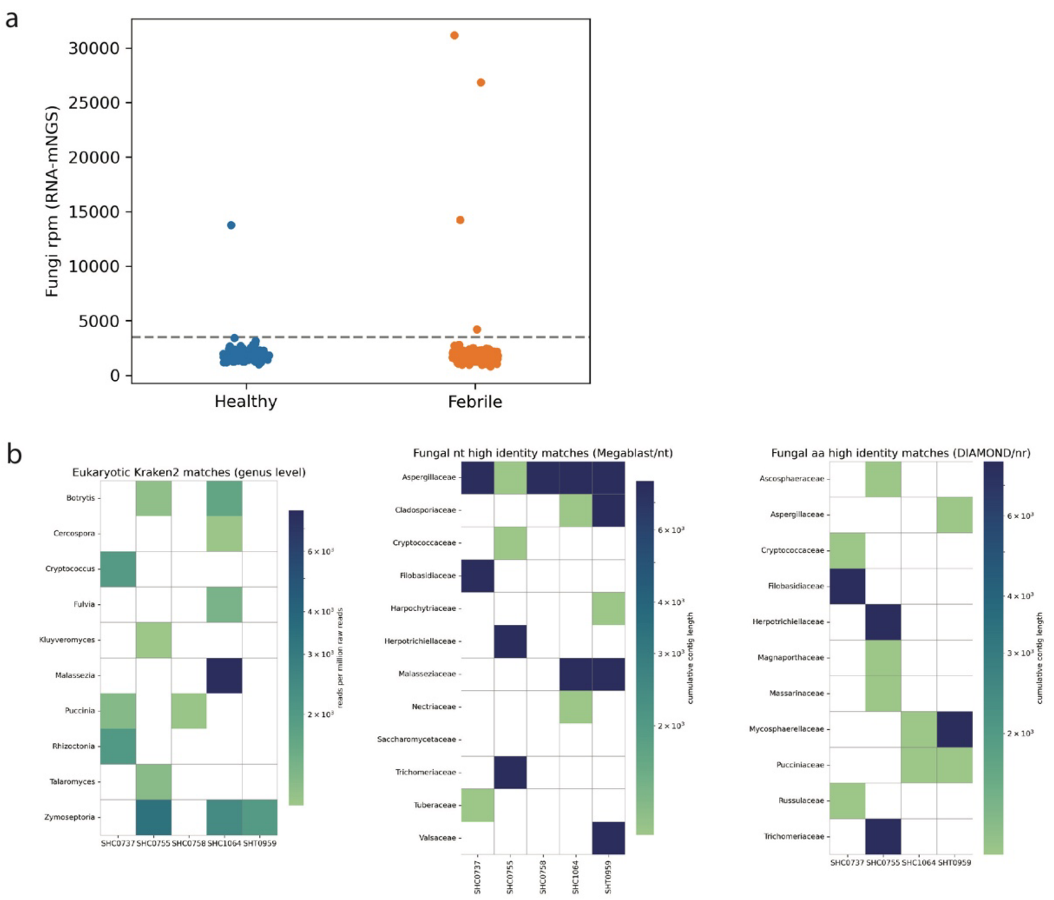


**Supplementary Figure 8: a.** Proportion of reads classified as fungal (reads per million raw reads, Kraken2) in febrile cases and healthy controls. **b.** Taxonomic classifaction (Kraken2) of fungal reads; color indicates reads per million, filtered to show only samples/genera with >1000 rpm. **c.** Fungal de novo contigs (SPAdes) classified by nucleotide search (megablast / nt) with > 90% sequence identity and >30% query coverage, filtered to show only samples/families with cumulative contig length > 1kb. **d.** Fungal de novo contigs (SPAdes) classified by translated nucleic acid search (DIAMOND-blastx / nr) with > 90% sequence identity and >30% query coverage, filtered to show only samples/families with cumulative contig length > 1kb.

a

| *Model type:* | **Clinical only** | **Clinical only** | **Clinical only** | **Clinical + CBC** |
|---|---|---|---|---|
| *Comparison group(s):* | **All febrile** | **Other NMFI** | **Other NMFI** | **Other NMFI** |
| *Training set:* | **2018-2019 (n = 526)** | **2018 (n = 288)** | **2019 (n = 163)** | **2019 (n = 163)** |
| *Testing set:* | **2018-2019 (n = 526)** | **2019 (n = 163)** | **2019 (n = 163)** | **2019 (n = 163)** |
| **Recall (95% CI)** | 0.922 (0.909-0.944) | 0.832 (0.797-0.889) | 0.815 (0.710-0.885) | 0.853 (0.769 – 0.909) |
| **Precision (95% CI)** | 0.847 (0.806-0.893) | 0.776 (0.730-0.878) | 0.773 (0.625 – 0.833) | 0.799 (0.767-0.833) |
| **F1 (95% CI)** | 0.883 (0.857-0.902) | 0.802 (0.774-0.854) | 0.793 (0.678-0.853) | 0.825 (0.769 – 0.870) |
| **AUC/ROC (95% CI)** | 0.921 (0.911-0.937) | 0.864 (0.821-0.920) | 0.871 (0.808-0.934) | 0.867 (0.842-0.895) |
| **AUC/PR (95% CI)** | 0.918 (0.888-0.956) | 0.843 (0.757-0.931) | 0.871 (0.819-0.923) | 0.828 (0.760-0.898) |

b Clinical only, all febrile, 2018-2019

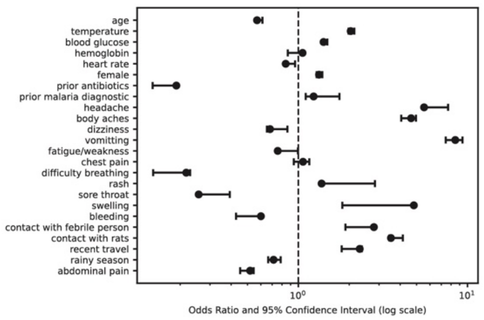


c Clinical only, Other NMFI, train 2018, test 2019

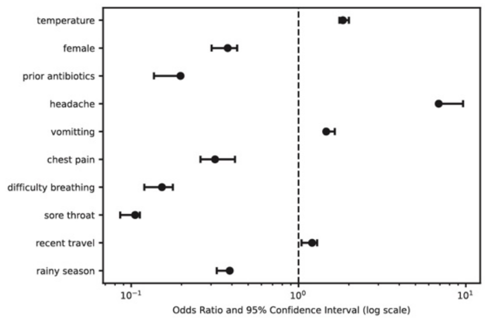


d Clinical only, Other NMFI, train 2019, test 2019

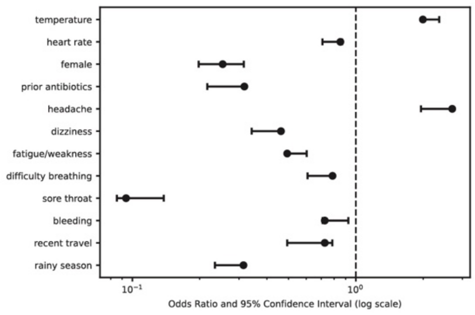


e Clinical + CBC, Other NMFI, train 2019, test 2019

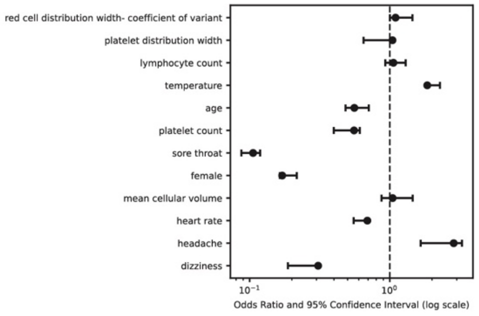


**Supplementary Figure 9: a.** Performance (tested with bootstrapping with 5-fold cross validation) of weighted logisistic regression models to distinguish *Borrelia* infection from all other febrile illness (All febrile) or all non-malarial febrile illness (Other NMFI) using clinical data, including demographics, symptoms, exposures, and vital signs (Clinical only) or clinical data and compelte blood counts with differential (Clinical + CBC). Odds ratios with 95% CI shown for model features in **b.** the clinical only model to distuinguish *Borrelia* from all other febrile, trained and tested on the full dataset from 2018-2019 (n = 526), **c.** the clinical only model to distuinguish *Borrelia* from all non-malarial febrile illness, trained on the 2018 dataset (n = 288) and tested on the 2019 dataset (n = 163), **d.** the clinical only model to distuinguish *Borrelia* from all non-malarial febrile illness, trained and tested on the 2019 dataset (n = 163), and **e.** the clinical and CBC model to distuinguish *Borrelia* from all non-malarial febrile illness, trained and tested on the 2019 dataset (n = 163).
